# Atypia of Undetermined Significance and ThyroSeq v3 Positive Call Rates as Quality Control Metrics for Cytology Laboratory Performance

**DOI:** 10.1101/2024.01.24.24301631

**Authors:** Odille Mejia-Mejia, Andres Bravo-Gonzalez, Monica Sanchez-Avila, Youley Tjendra, Rodrigo Santoscoy, Katherine Drews-Elger, Yiqin Zuo, Camilo Arias-Abad, Carmen Gomez, Monica Garcia-Buitrago, Mehrdad Nadji, Merce Jorda, Jaylou M. Velez-Torres, Roberto Ruiz-Cordero

## Abstract

**Background:** The Bethesda system (TBS) for reporting thyroid cytopathology recommends an “atypia of undetermined significance (AUS)” rate of 10%. Recent data suggest that this category might be overused when the rate of cases with molecular positive results is low. As a quality metric, we calculated the AUS and positive call rates for our cytology lab and each cytopathologist.

**Methods:** A retrospective analysis of all thyroid cytology cases in a 4.5-year period was performed. Cases were stratified by TBS category, and molecular testing results were collected for indeterminate categories. The AUS rate was calculated for each cytopathologist (CP) and the laboratory. The molecular positive call rate (PCR) was calculated with and without the addition of *currently negative* to the *positive* results obtained from the ThyroSeq report.

**Results:** 7,535 cases were classified as *non-diagnostic* 7.6%, *benign* 69%, *AUS* 17.5%, *follicular neoplasm / suspicious for follicular neoplasm* 1.4%, *suspicious for malignancy* 0.7 %, and *malignant* 3.8%. The AUS rate for each cytopathologist ranged from 9.9-36.8%. The overall PCR for the cytology laboratory was 24% (range 13-35.6% per CP). When including cases with *currently negative* results, the PCR increased to 35.5% for the cytology laboratory (range 13-42.6% per CP). Comparison analysis indicates a combination of overcalling benign cases and, less frequently, under calling of higher TBS category cases.

**Conclusions:** The AUS rate in the context of PCR is a useful metric to assess cytology laboratory and cytopathologists’ performance. Continuous feedback on this metric could help improve the overall quality of reporting thyroid cytology.

## Introduction

Thyroid nodules are common and can be seen in approximately half of the population above 60 years of age ^1^. While most nodules are benign, certain clinical and/or sonographic criteria may raise concern for malignancy and lead to additional exams, including microscopic examination of the cells via fine needle aspiration cytology (FNAC) ^2,3^. In this context, cytomorphologic evaluation serves as a surrogate to infer underlying genomic alterations that can drive tumorigenesis and dictate the malignant potential of the nodule.

The Bethesda System for Reporting Thyroid Cytopathology (TBSRTC) proposes six diagnostic categories associated with different risk of malignancy (ROM) based on FNAC findings, guiding further management^1^. Among the indeterminate categories (i.e., atypia of undetermined significance/follicular lesion of undetermined significance [AUS/FLUS, TBSRTC III], follicular neoplasm or suspicious for follicular neoplasm [TBSRTC IV], and suspicious for malignancy [TBSRTC V]), TBSRTC advises an AUS rate of no more than 10% and recommends the use of molecular testing to better stratify these nodules. The relevance of molecular testing in thyroid cytology has been reflected in the most recent 3^rd^ edition of TBSRTC^2^.

While molecular testing is not the gold standard for the diagnosis of thyroid nodules and has limitations, its use has been proven beneficial not only in avoiding unnecessary surgeries but also in impacting clinical management, selection of targeted therapy for some malignant cases, and, more recently, as a quality control metric among cytopathologists (CPs) and the cytology laboratory ^3,4^.

As cytopathologic interpretation of thyroid nodules informs on treatment decisions and impacts patient outcomes, accurate diagnosis is essential for guiding patient care. To this end, assessing cytopathologist performance is crucial.

In this study, we demonstrate that the AUS and molecular positive call rates (PCR) are useful quality control metrics to assess performance, both on an individual cytopathologist basis, as well as for the cytology laboratory. Moreover, by introducing time as a variable, we identified significant trends that offer more objective and valuable feedback to each CP.

## Methods

Under Institutional Review Board approval from the University of Miami, we retrieved retrospective data on all thyroid FNACs performed and evaluated at the University of Miami Hospital between January 2018 to July 2022. Each FNAC was executed under ultrasound guidance using 25G needles by interventional radiologists or endocrinologists without cytology-assisted rapid on-site adequacy assessment. Direct conventional smears fixed in ethanol and needle rinses and/or direct passes collected into CytoLyt solution for preparing a ThinPrep (Hologic, Malborough, MA) slide were delivered to the cytology laboratory and stained with Papanicolaou stain. Additionally, needle rinses and/or direct passes were collected into the provided media vial for molecular testing. The biopsy results were evaluated and reported by one of 10 board-certified CPs according to TBSRTC second edition criteria^1^. Data collection was extracted from our laboratory information system and included the date of collection, reporting CP, patient’s age, gender, anatomic location of the FNAC, diagnosis, and TBSRTC category.

Descriptive statistics such as frequencies and percentages were computed for categorical variables to provide a clear overview of their distribution. On the other hand, non-categorical variables underwent continuous data analyses. Descriptive statistics such as means, medians, standard deviations, and ranges were computed to summarize the central tendencies and variabilities of these variables, as appropriate. A Chi-squared test or Student’s *t*-test were used for comparing distributions of categorical data or the means between two groups, as appropriate. Statistical analyses and visualization plots were conducted using the R programming language (R Core Team, 2023). Statistical significance was established as a p-value of less than 0.05.

### TBSRTC Rates

We use standardized canned comments for reporting thyroid FNACs using TBSRTC 2017 2^nd^ edition, which facilitate the consistency and extraction of data^5^. The utilization rates of TBSRTC categories were determined by dividing the number of cases within each category by the total number of cases annually and throughout the entire study period, both for the laboratory and for each CP. The rates are expressed in percentages.

### Molecular Positive Call Rate (PCR)

Per institutional protocol, nodules classified as AUS/FLUS, FN/SFN, and, more recently, suspicious for malignancy are reflexed to molecular testing. In 2018, candidate samples were sent to either ThyroSeq V3 (Sonic Healthcare, Rye Brook, NY) or Afirma (Veracyte, South San Francisco, CA) for molecular testing. From 2019 onwards, all samples were sent exclusively to ThyroSeq. Specific molecular alterations, including sample adequacy, results, probability of malignancy, gene mutated, specific mutation(s), allelic frequency, copy number alterations, fusions, and gene expression, were manually copied from the reports. For cases classified as AUS/FLUS that underwent molecular testing, the PCR was calculated by dividing the cases with *positive* ThyroSeq results by the total number of AUS cases that underwent molecular testing^6^.

Because molecular alterations, even when not directly associated with increased malignant potential, can instigate cytomorphologic alterations, and because current recommendations from the National Comprehensive Cancer Network (NCCN) guidelines suggest active surveillance for neoplastic nodules with a low risk of malignancy^7,8^, we also conducted an analysis that included cases with *positive* and *currently negative* ThyroSeq results in the calculation of the PCR to investigate whether incorporating such cases would yield a significant difference in our PCR calculations. Rates with and without *currently negative* cases were statistically compared using a Z-test for two proportions to assess differences by CP and for the entire laboratory.

A dedicated dashboard for enhanced visualization and analysis of the results is available: http://impossiblecode.pythonanywhere.com/.

## Results

### Cohort Characteristics

Between January 2018 and July 2022, a comprehensive dataset of thyroid nodule FNACs was collected. This dataset comprised a total of 7,535 FNAC specimens thoroughly examined by a team of 10 CPs. The workload distribution among CPs varied significantly, with individual case counts for the study period ranging from 57 to 1,802 (Figure 1 and Table 1). Within this cohort, the patient demographic landscape was diverse. Age exhibited a normal distribution for both sexes, with a median age of 57 years and a range of 14 to 95 years. As expected, most of the cases, accounting for 81.2% (6,120 cases), pertained to female patients. Across the different years evaluated, there were 1,562 cases in 2018, constituting 20.7% of the dataset. The following year, 2019, contributed 1,753 cases, representing 23.3% of the cohort. In 2020, there were 1,551 cases (20.6%), while 2021 added 1,618 cases (21.5%). The first half of 2022 accounted for 1,051 cases (13.9%) (Table 1).

**Fig 1.**
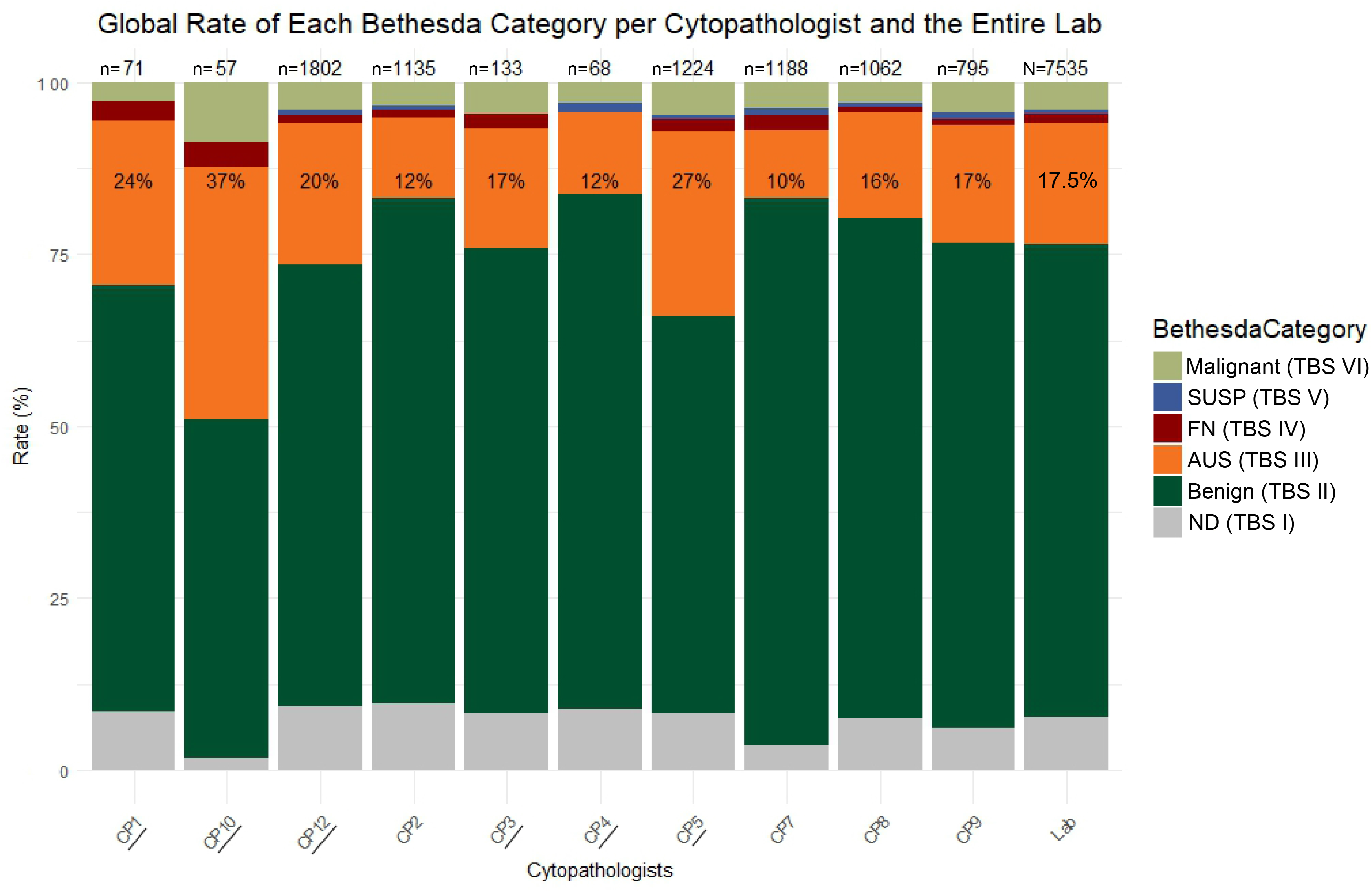
Stacked bar graph showing the distribution of thyroid FNA interpretations based on TBS diagnostic category stratified by individual cytopathologists (CP). Total number of cases interpreted by each CP appear atop the bars. TBS = the Bethesda System.

**Table 1.**
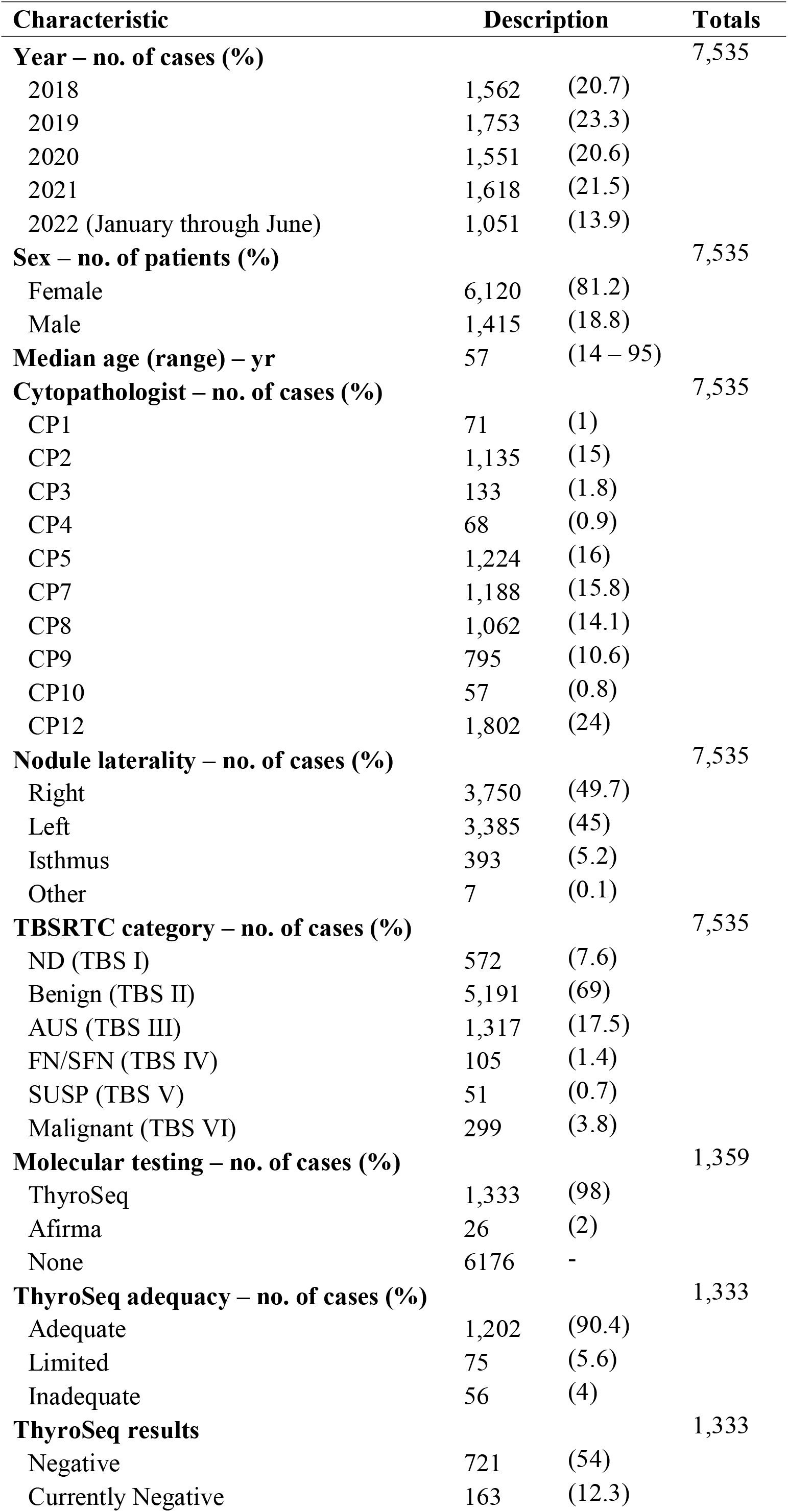
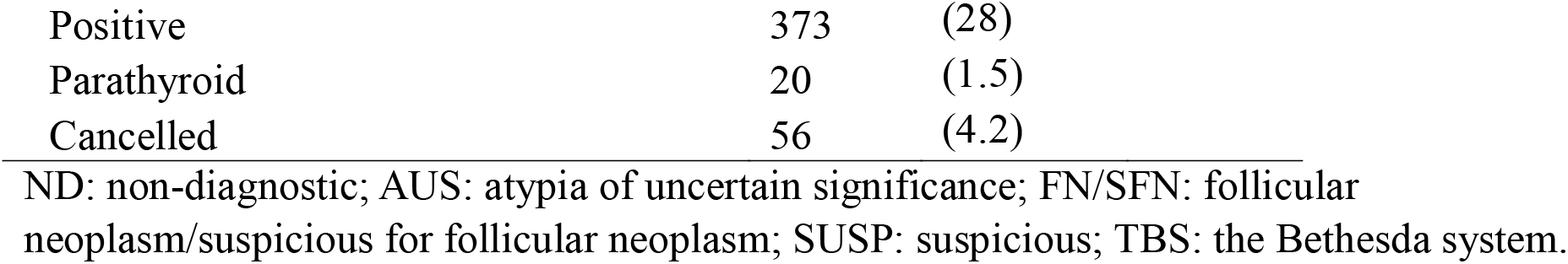
Summary Statistics of the Study Cohort.

| Characteristic | Description |  | Totals |
| --- | --- | --- | --- |
| **Year – no. of cases (%)** |  |  | 7,535 |
| 2018 | 1,562 | (20.7) |  |
| 2019 | 1,753 | (23.3) |  |
| 2020 | 1,551 | (20.6) |  |
| 2021 | 1,618 | (21.5) |  |
| 2022 (January through June) | 1,051 | (13.9) |  |
| **Sex – no. of patients (%)** |  |  | 7,535 |
| Female | 6,120 | (81.2) |  |
| Male | 1,415 | (18.8) |  |
| **Median age (range) – yr** | 57 | (14 – 95) |  |
| **Cytopathologist – no. of cases (%)** |  |  | 7,535 |
| CP1 | 71 | (1) |  |
| CP2 | 1,135 | (15) |  |
| CP3 | 133 | (1.8) |  |
| CP4 | 68 | (0.9) |  |
| CP5 | 1,224 | (16) |  |
| CP7 | 1,188 | (15.8) |  |
| CP8 | 1,062 | (14.1) |  |
| CP9 | 795 | (10.6) |  |
| CP10 | 57 | (0.8) |  |
| CP12 | 1,802 | (24) |  |
| **Nodule laterality – no. of cases (%)** |  |  | 7,535 |
| Right | 3,750 | (49.7) |  |
| Left | 3,385 | (45) |  |
| Isthmus | 393 | (5.2) |  |
| Other | 7 | (0.1) |  |
| **TBSRTC category – no. of cases (%)** |  |  | 7,535 |
| ND (TBS I) | 572 | (7.6) |  |
| Benign (TBS II) | 5,191 | (69) |  |
| AUS (TBS III) | 1,317 | (17.5) |  |
| FN/SFN (TBS IV) | 105 | (1.4) |  |
| SUSP (TBS V) | 51 | (0.7) |  |
| Malignant (TBS VI) | 299 | (3.8) |  |
| **Molecular testing – no. of cases (%)** |  |  | 1,359 |
| ThyroSeq | 1,333 | (98) |  |
| Afirma | 26 | (2) |  |
| None | 6176 | - |  |
| **ThyroSeq adequacy – no. of cases (%)** |  |  | 1,333 |
| Adequate | 1,202 | (90.4) |  |
| Limited | 75 | (5.6) |  |
| Inadequate | 56 | (4) |  |
| **ThyroSeq results** |  |  | 1,333 |
| Negative | 721 | (54) |  |
| Currently Negative | 163 | (12.3) |  |

|  |  |  |
| --- | --- | --- |
| Positive | 373 | (28) |
| Parathyroid | 20 | (1.5) |
| Cancelled | 56 | (4.2) |

ND: non-diagnostic; AUS: atypia of uncertain significance; FN/SFN: follicular neoplasm/suspicious for follicular neoplasm; SUSP: suspicious; TBS: the Bethesda system.

Notably, four CPs (CP1, CP3, CP4, and CP10) only signed out 329 cases, accounting for 4.3% of the entire cohort (individual range: 57 to 133 cases). This variation in caseload distribution may lead to over or underrepresentation of diagnostic category rates for these CPs but has minimal impact on the overall laboratory rates, as demonstrated by a Chi-square test showing a p-value of 1.0.

### Bethesda Category Rates – Global and Over Time

#### The Cytology Laboratory

TBSRTC rates for the study period show a *non-diagnostic* rate of 7.6%, a *benign* rate of 69%, an *AUS* rate of 17.5%, a *follicular lesion/neoplasm* rate of 1.4%, a *suspicious for malignancy* rate of 0.7 %, and a *malignant* rate of 3.8% (Table 1, Figures 1 and 2). Overall, the AUS rate of the laboratory during the 4.5 years examined was slightly higher than the recommended 10%. When plotted over time, the AUS rates showed a steady decline, starting with 17.8% in 2018 to 15.5% in 2021; however, for the first half of 2022, the AUS rate increased by 6% to 21.5% (Figure 3). Notably, all CPs, except for CP5, showed an increase in the use of the AUS category, which explains the overall marked increase (Figure 4).

**Fig 2.**
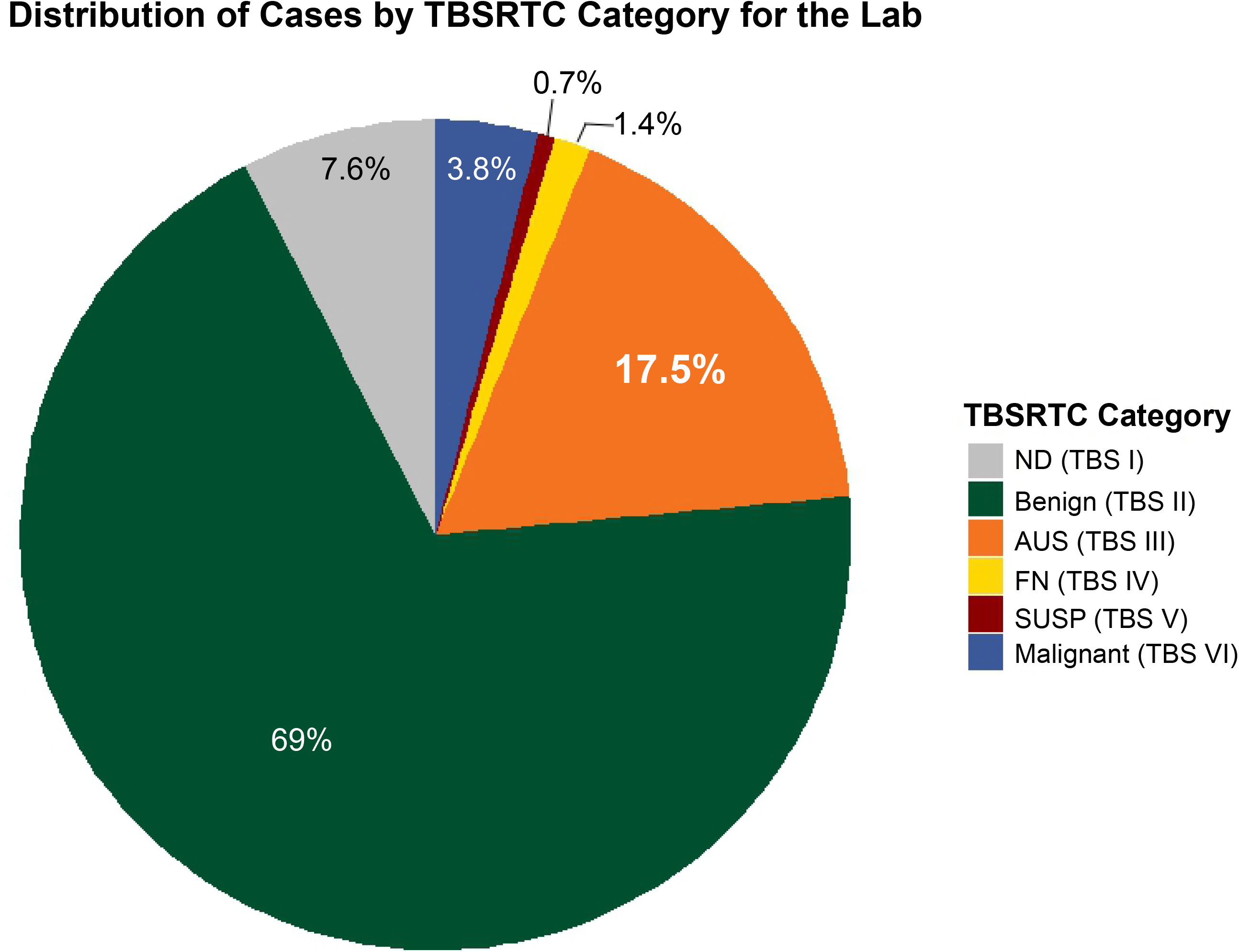
Pie chart showing the proportion of cases based on the Bethesda System categories.

**Fig 3.**
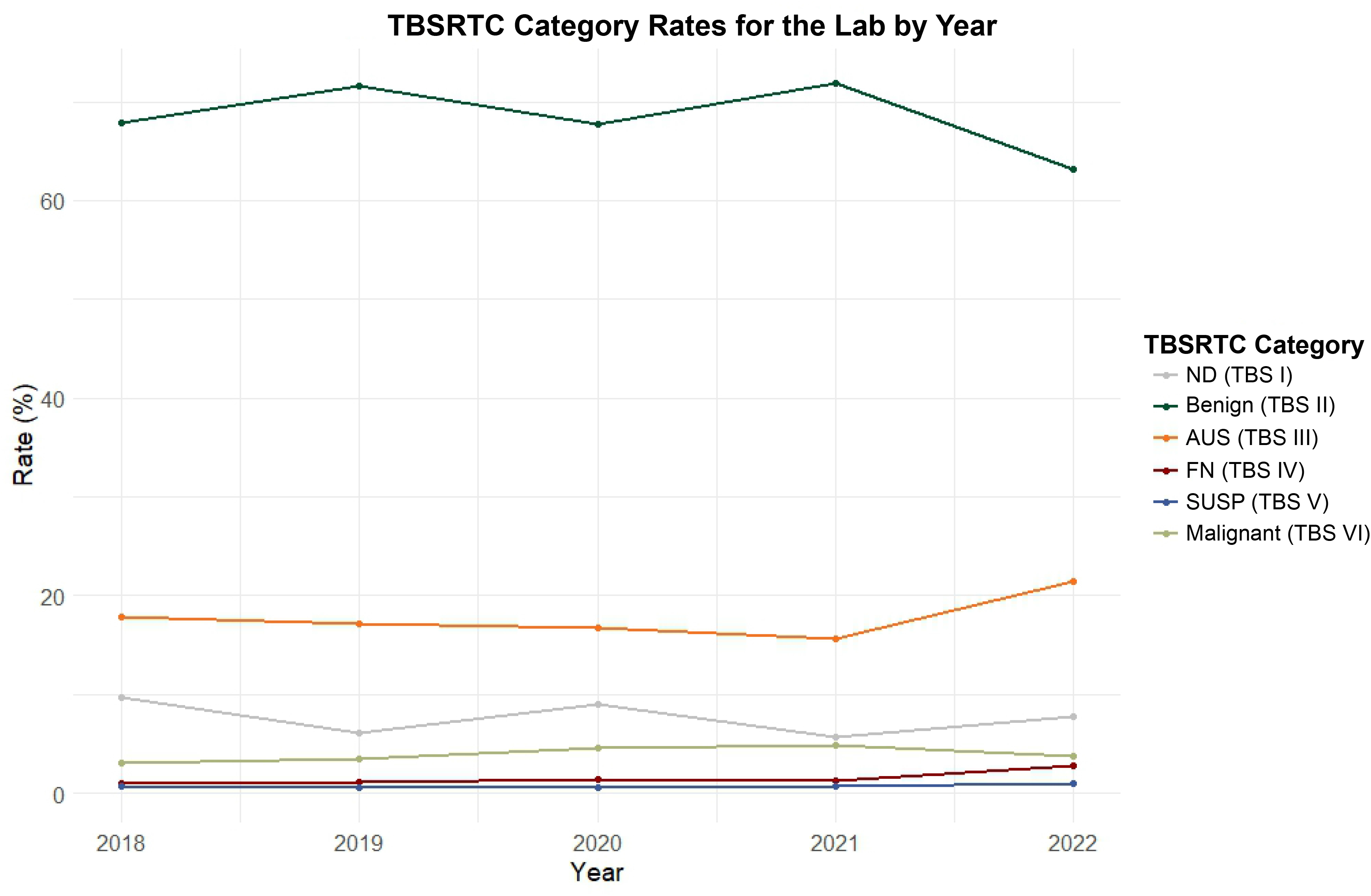
Line plot showing the proportion of the Bethesda System categories for the laboratory per year.

**Fig 4.**
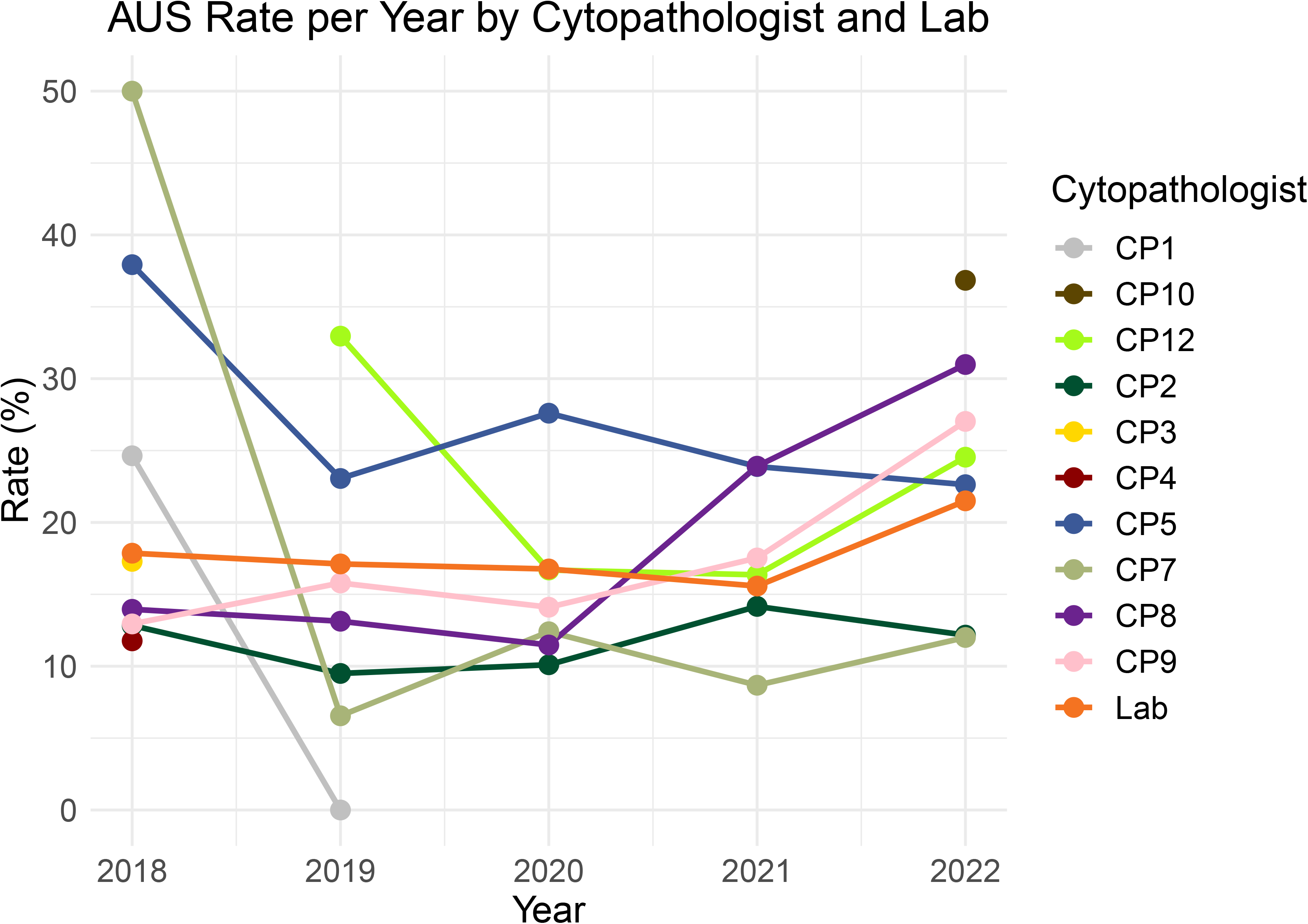
Line plot showing the AUS rate by each cytopathologist per year. Cytopathologist with one year worth of data appear as a single dot.

#### Individual CPs Rates

Individual CPs’ rates showed a variable distribution, with some CP’s rates resembling the overall laboratory rates, while others exhibiting significant differences (Figure 1). Over the 4.5-year study period, the AUS rate per CP fluctuated between 9.93% and 36.8% and showed significant variability over time (Figure 4). This variability was partly influenced by the cumulative number of cases each CP handled.

### Molecular Testing Results of AUS Cases

A total of 1,473 cases were classified under TBS categories III (1,317, 89.4%), IV (105, 7.3%), and V (51, 3.3%). Based on the ThyroSeq reports, *adequate/limited* nucleic acid extraction was sufficient to perform sequencing in 1,277 (96%) cases and *inadequate* in 56 (4%) cases. Only 26 (2%) cases were sent to Afirma in 2018 and were excluded from subsequent analyses (Table 1).

The ThyroSeq report summarizes the findings into *negative*, *currently negative*, *positive*, or presence of *parathyroid* or *C-cells/medullary carcinoma*. Of the 1,317 AUS cases in our cohort, 1,226 (93%) underwent ThyroSeq testing. Sequencing results were *negative* in 721 (55.8%), c*urrently negative* in 163 (12.3%), and *positive* in 373 (28%) cases. The most frequent molecular alterations at the gene level of *currently negative* and *positive* cases (534) included mutations in the RAS gene family (*NRAS, KRAS*, and *HRAS*) observed in 39% (208) of cases, followed by *TSHR* mutations in 13% (70) of cases, and *BRAF* alterations in 10% (54) of cases (Figure 5). Of note, *TERT* and *TP53* were mutated in 12 (2.2%) and 9 (1.6%) cases, respectively. With regards to gene fusions, 39 cases (7%) showed activating fusions involving genes *PAX8* (13, 2.4%), *THADA* (11, 2%), *RET* (6, 1.1%), *NTRK3* (4, 0.7%), *ALK* (2, 0.3%), and *BRAF* (2, 0.3%) (Figure 5). The most frequent gene implicated in the *currently negative* cases was *TSHR,* followed by *EIF1AX* (23, 4.3%), *EZH1* (16, 2.9%), and *PTEN* (9, 1.6%) (Figure 6). In a limited number of cases (23, 4.3%), RNA gene expression profiling showed overexpression of the NIS (*SLC5A5*) gene. This finding is indicative of an autonomous functional nodule but does not indicate a neoplastic process, for this reason, these cases were considered as *negative.* Notably, we observed a statistically significant difference in age across different mutated genes (Supplement Figure 2). Mutations in genes *EIF1AX*, *GNAS*, and *EZH1* were more frequently observed in older patients (median age 60 years), whereas mutations in genes *DICER1* (median age 41.5 years) and *NRAS* (median age 48.5 years) were more frequently seen in patients under 50 years of age (p = 0.0002).

**Fig 5.**
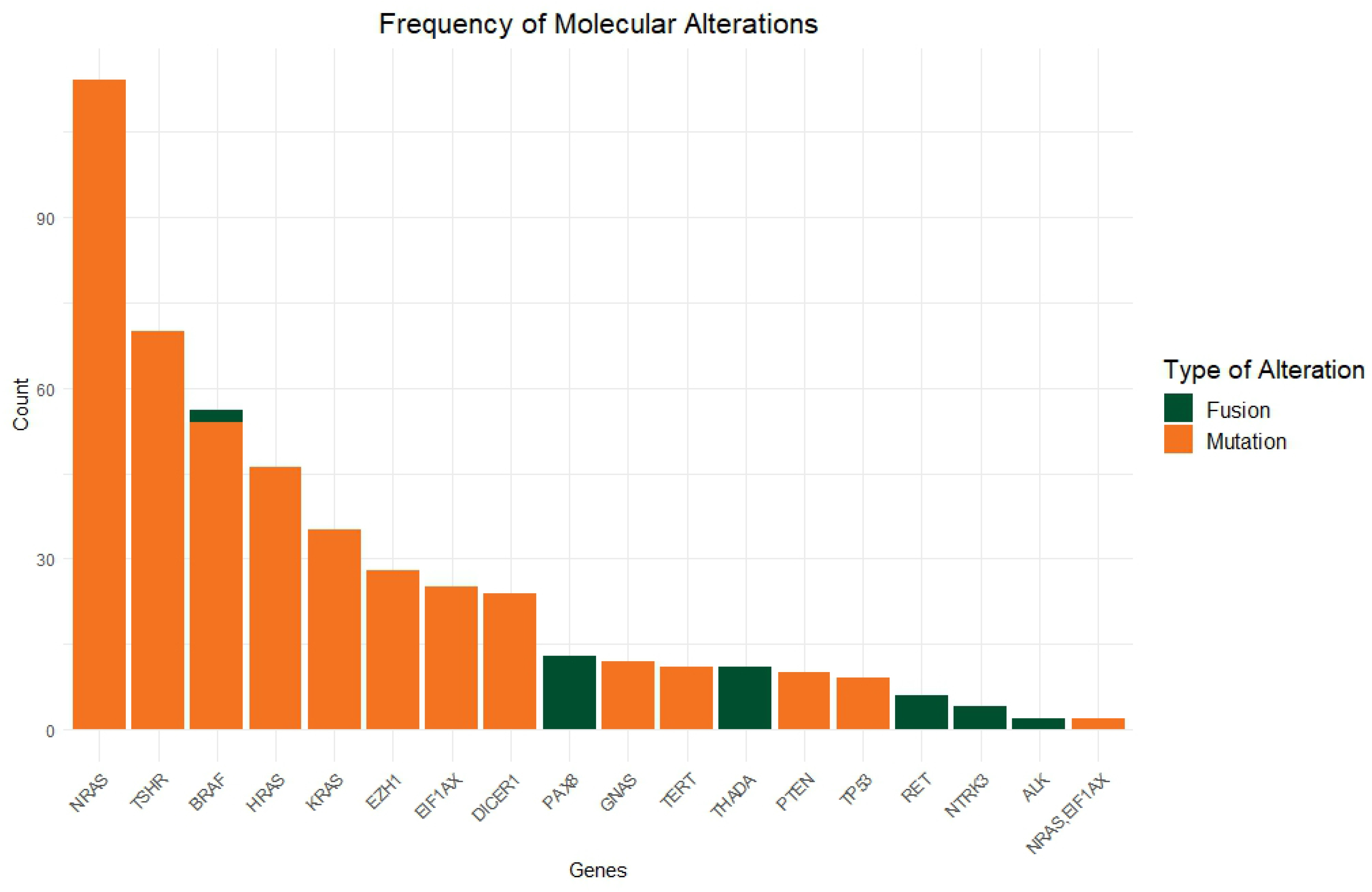
Absolute frequency of molecular alterations based on ThyroSeq V3 results.

**Fig 6.**
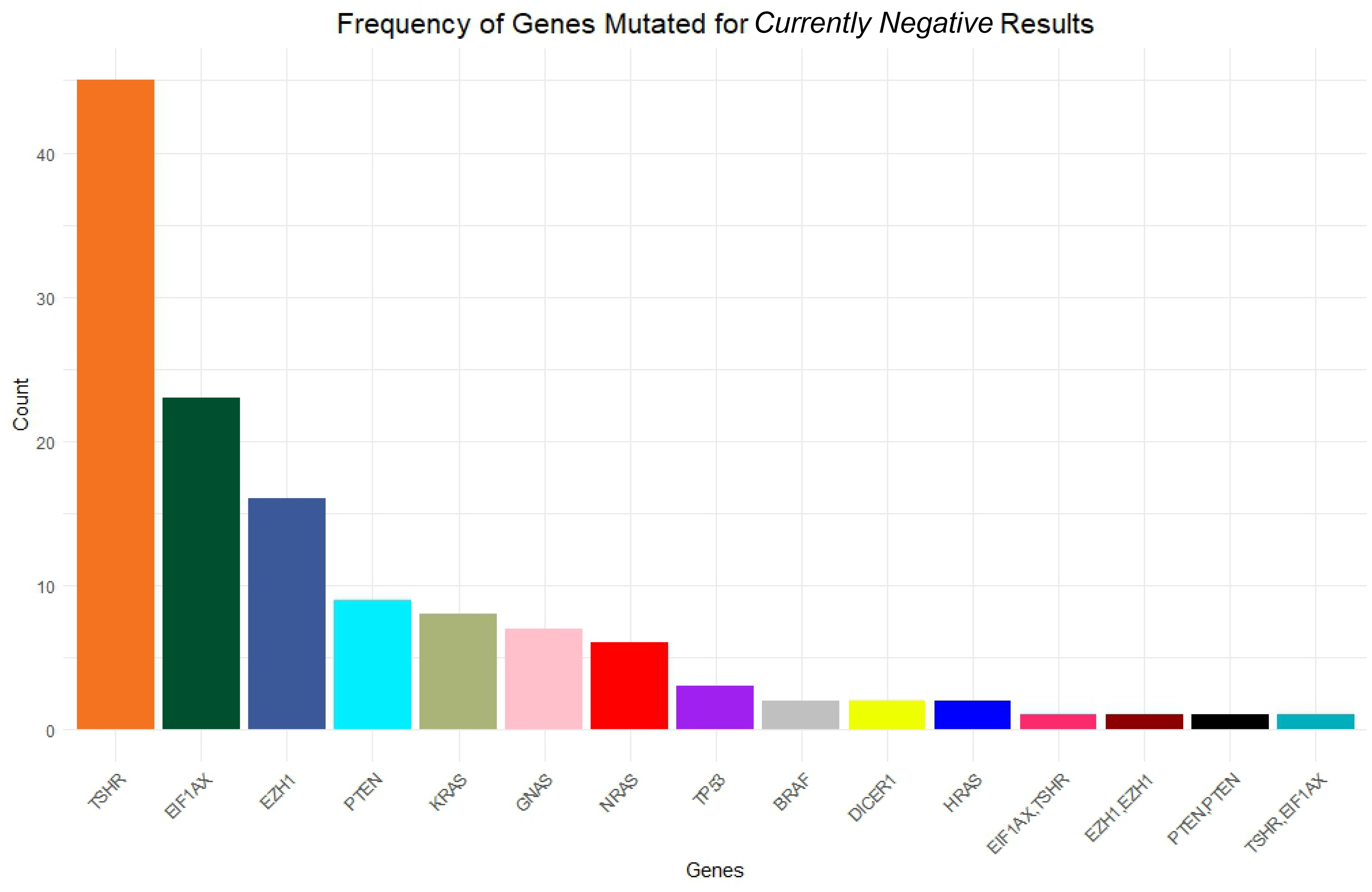
Absolute frequency of gene mutations described in samples classified as "currently negative" by ThyroSeq V3 results.

### Positive Call Rate – Global and Over Time

#### The Cytopathology Laboratory

As a whole, the cytopathology laboratory showed a PCR for AUS cases of 24% when including only cases with *positive* ThyroSeq results, and 35.5% when including cases with *positive* and *currently negative* results (p < 0.0001) (Table 2 and Figure 7). When analyzed over time, the inclusion of *currently negative* cases led to an increase in the PCR for most years (Supplement Figure 1).

**Fig 7.**
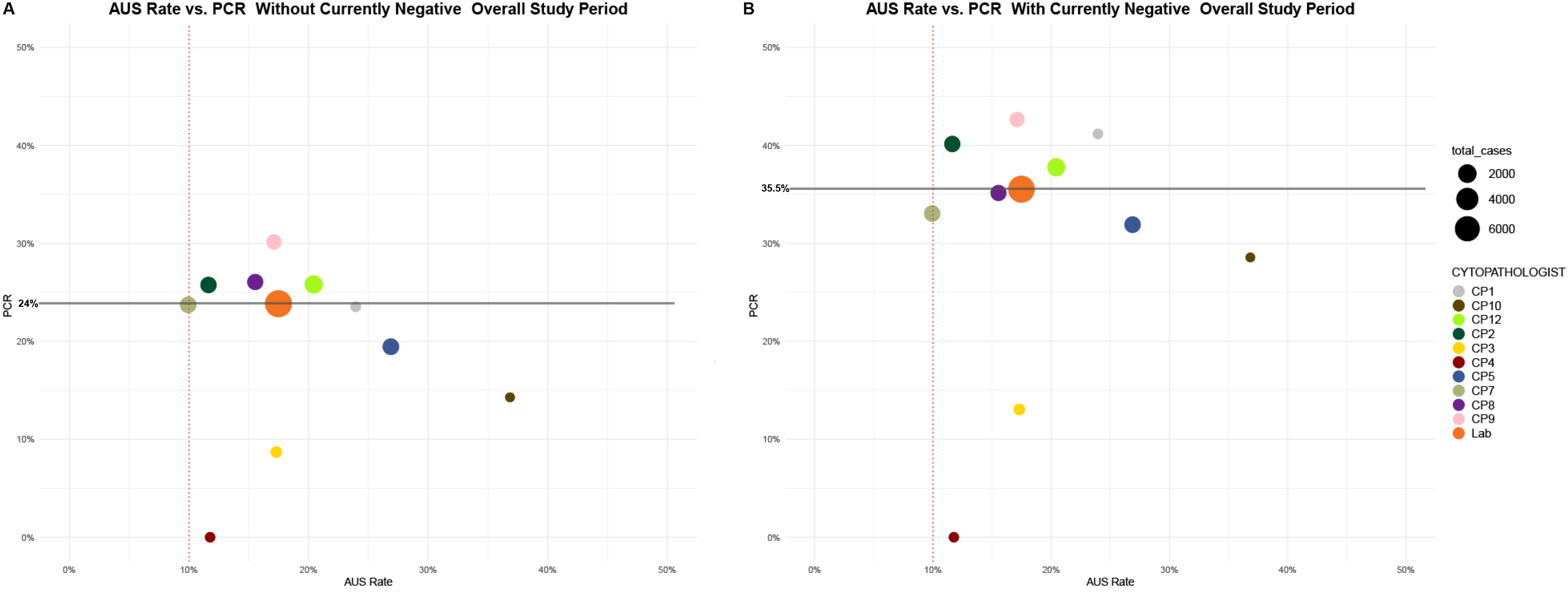
**A)** Scatterplot showing AUS rate vs PCR for the entire study period calculated without *currently negative* cases. **B)** Scatterplot showing AUS rate vs PCR for the entire study period calculated including the *currently negative* cases. Each bullet represents a different cytopathologist. The size of the bullet is proportional to the number of cases evaluated. The large orange bullet represents the cytopathology laboratory. The vertical dotted red lines indicate the recommended 10% AUS rate. The solid-grey horizontal lines indicate the PCR of the laboratory.

**Table 2.** AUS Rate and PCR by Cytopathologist.

| Cytopathologist | Total #<br>of AUS cases | AUS Rate<br>(%) | PCR without<br>*Currently Negative* (%) | PCR with<br>*Currently Negative* (%) | p-value |
| --- | --- | --- | --- | --- | --- |
| CP1 | 17 | 24 | 24 | 41 | 0.3 |
| CP10 | 21 | 37 | 14 | 28 | 0.26 |
| CP12 | 368 | 20 | 26 | 38 | 0.0004* |
| CP2 | 132 | 11 | 26 | 40 | 0.01* |
| CP3 | 23 | 17 | 9 | 13 | 0.63 |
| CP4 | 8 | 12 | 0 | 0 | NA |
| CP5 | 329 | 27 | 20 | 32 | 0.0002* |
| CP7 | 118 | 9 | 24 | 33 | 0.11 |
| CP8 | 165 | 15 | 26 | 35 | 0.07 |
| CP9 | 136 | 17 | 30 | 43 | 0.03* |
| Lab | 1,317 | 17.5 | 24 | 35 | <0.0001* |

CP = cytopathologist; AUS = atypia of uncertain significance; PCR = positive call rate. P-value indicates the Z-test for two proportions between PCR with and without *currently negative*, to assess differences by cytopathologist and for the entire cytopathology laboratory. \* Indicates statistical significance.

#### Individual CPs Rates

Individual assessment of the global PCR yielded interesting results by CPs. For four CPs, the difference in PCR with and without *currently negative* was statistically significant (p<0.05), while for the rest, it did not reach statistical significance (Table 2). When assessed over time, some CPs show relatively stable rates, and others show wider variability, partly influenced by the total number of cases examined (Supplement Figure 1).

### AUS Rate in the Context of the PCR

While calculating the AUS rate provides insights into the usage of this TBS category concerning the recommended 10% threshold, it alone cannot pinpoint the root cause of its overutilization (overcalling benign nodules vs. undercalling neoplastic nodules). However, when we plot the AUS rate against the PCR, it becomes feasible to discern whether the overutilization stems from an excessive diagnosis of benign (TBS II) nodules, as indicated by a high AUS rate (>10%) and low PCR, or from an underdiagnosis of TBS IV, V, or VI category nodules, seen in cases with high AUS rate (>10%) and high PCR (Figure 7). This comprehensive analysis for the entire laboratory encapsulates both scenarios, which can only be revealed by calculating the AUS rate and PCR for each CP. Using the laboratory PCR as a benchmark is useful when evaluating individual PCRs.

## Discussion

Our study demonstrates that retrospective assessment of the AUS rate in the context of PCR, obtained via ThyroSeq V3 results, is a valuable metric for evaluating CP and overall laboratory performance.

In our laboratory, the global AUS rate for the study period was 17.5%, slightly higher than the recommended 10% rate by TBS ^1^, but still within the range reported by other laboratories in the literature. For example, AUS rates as low as 4.5% have been reported by Al-Abbadi et al. ^9–12^, whereas Bernstein et al. found a rate of 12% ^13^, Hathi et al. showed a similar rate to ours of 18.8% ^14^, and Wu et al. showed a rate of 27.2% ^15^.

We individualized AUS rates by CPs, as depicted in Figure 1. This approach has been proposed to improve the accuracy and consistency of cytology evaluation because it provides real-time data-driven feedback to CPs, enabling them to closely monitor their proximity to the AUS rate threshold and refine their classification process accordingly. Horback et al. developed a thyroid and cervical cytology dashboard, which displayed individual CP reporting rates in different zones based on their proximity to the benchmark ^16^. Similarly, Vanderlaan et al. developed a scheme for thyroid cytology smears and found that most CPs were not meeting the established limit in their laboratory ^4^. These approaches are being studied to reduce inter-observer variability and improve the accuracy of cytology evaluation. It is important to remember that rate variations can be attributed to various factors, such as the types of patients who seek care, the number of samples evaluated, and the experience of the CPs evaluating the samples, among others. However, despite the variability in reported rates, we should still strive to achieve the recommendation proposed by TBSRTC. Of note, it is critical to emphasize that merely reducing AUS rates without recalibrating individual diagnostic thresholds could potentially impair the diagnostic sensitivity of thyroid FNAC. Therefore, to maintain a balance between lowering AUS rates and preserving diagnostic accuracy, we have implemented a dashboard to track the utilization of AUS and PCR and have established a monthly conference to review the cytomorphology of thyroid FNAC with *positive* and *currently negative* results. The impact of these interventions is unclear at this point and will be the subject of future studies.

Molecular diagnosis has emerged as a promising approach for stratifying patients with AUS nodules, helping to avoid more invasive diagnostic procedures. In a recent meta-analysis, three diagnostic tools - Afirma, ThyroSeq V2, and ThyroSeq V3 - were compared, and it was found that ThyroSeq v3 had the highest performance with an area under the curve (AUC) of 0.95 (confidence interval of 0.93 - 0.97), followed by Afirma (AUC of 0.90) and ThyroSeq V2 (AUC of 0.88) ^17^. In our study, the *benign call rate* (BCR) was 55.8% when considering only the cases with *negative* ThyroSeq results and 68.1% when including cases with *negative* and *currently negative* results. These rates are comparable with those reported by Steward et al. (61%)^18^ and Desai et al. (71%) ^19^, who also included *currently negatives* in the BCR. While it has been documented that a high BCR translates into an increased number of patients for whom more invasive procedures are spared, currently, there is no recommendation as to what the target BCR or PCR should be. We believe that a high BCR also translates into overcalling benign nodules and abusing the use of the AUS category since a BCR rate of 90%, for example, would indicate that nine out of every ten AUS cases submitted for molecular testing yield a *negative* result.

On the contrary, a very low BCR (∼10%) would suggest undercalling cases that should be better classified as IV, V, or even VI. Based on our analysis, it is unclear what the optimal PCR should be, and additional studies taking into consideration the prevalence of thyroid pathologies in a given population and the total number of cases reviewed per CP are necessary to make this determination. In the meantime, as suggested by Vanderlaan et al.^4^, a good practice is to use the global PCR of the cytology laboratory as a benchmark.

The calculation of PCR with and without the inclusion of cases with *currently negative* ThyroSeq results offers valuable insights into both clinical management and CP performance assessment. Including cases with *currently negative* results in the PCR calculation provides a comprehensive view of the diagnostic landscape. From a clinical management perspective, this approach acknowledges the potential impact of molecular alterations on patient care. It recognizes that certain genetic changes, while not indicative of immediate malignancy, can still influence clinical decisions as indicated in the NCCN guidelines. ^7^. From a pure performance assessment approach, including *currently negative* cases in the calculation provides a clearer picture of how well CPs can identify cases with genetic alterations, which is crucial for evaluating their diagnostic skills.

Noteworthy, changes in the laboratory’s AUS rate affect the tests’ predictive values, and ThyroSeq v3 is no exception. Because sensitivity and specificity are constant values, the negative predictive value (NPV) and positive predictive value (PPV) can be affected by the prevalence of the disease in the population studied^18, 19^. As shown by Steward et al., having a disease prevalence over 40% in AUS would compromise the NPV of the molecular test, yielding suboptimal results ^18^. In our cohort, the disease prevalence by surgical thyroid specimen evaluation was not calculated for the entire cohort, but it is believed to be below this threshold since the PCR ranged from 24 to 35.5%^20^ depending on the exclusion or inclusion of *currently negative* cases, respectively.

The ROM associated with the AUS/FLUS category ranges ideally from 6-18%, but observational reports have found risks as high as 72.9% ^21^, suggesting that high AUS rates among CPs can be a negative quality indicator of diagnostic performance. If the CP is overestimating the observed malignancy features, the prevalence of thyroid cancer is expected to be lower. Still, if the CP underestimates malignancy, then, more TBS IV/V/VI are going to be misclassified as AUS, thus, potentially increasing the prevalence of thyroid cancer in the AUS population. The first case will decrease PPV, but the second case will affect the NPV^18^. A combination of overestimating and underestimating malignancy is also possible and frequently found ^4^. As shown by Ohori et al. ^22^ and confirmed by our group ^20^, the molecular-derived risk of malignancy (MDROM) approaches closely the true ROM and appears to be a better indicator of the malignant potential of a nodule.

In a similar study to ours, Vanderlaan et al. ^4^ assessed the PCR and AUS rates in their laboratory with results derived from Afirma molecular testing. Given that Afirma reports results as *"benign"* or ’’*suspicious"*, the PCR was calculated only taking the suspicious as positive with no ability to evaluate *currently negative* cases. They found an AUS rate of 22.3% and a PCR of 29% in a similar 3.5-year period (2018-2021). However, in this study temporality was not included as a metric and only global results were reported.

By tracking these metrics over time, we identified significant trends and practice changes, enabling us to offer more detailed feedback to CPs. For instance, within our cohort, CP5 initially exhibited a notably high AUS rate (40%) coupled with a low PCR (20%). Despite maintaining an elevated overall AUS rate of 27% throughout the study period, CP5 stands out as the only pathologist consistently demonstrating a year-over-year reduction in AUS rate (as depicted in Figures 1 and 4). Conversely, CP8 and CP9 have shown an increasing utilization of the AUS category over time, accompanied by a decreasing or stable PCR, respectively. While, on a global scale, these CPs exhibit better PCR and AUS rates compared to CP5, the introduction of temporal analysis reveals that CP5 is moving in the right direction, whereas CPs 8 and 9 appear to be increasing their reliance on the AUS category. We expect that by providing continuous feedback, our group will have a more homogeneous performance over time.

Genetic alterations were consistent with the current literature, showing a predominance of *RAS* gene mutations followed by *BRAF* and a minority of *TERT* and *TP53* alterations ^23^. The most common mutation in the *currently negative* category was *TSHR*, as expected since it is not indicative of malignancy but rather of neoplastic thyroid nodules with indolent behavior^24^.

The strengths of our study include extensive analysis of many cases over a 4.5-year span. The consistency in practice, particularly concerning the collection and dispatch of ThyroSeq, which is maintained due to adherence to institutional protocols. Nonetheless, there are discernible limitations such as the study’s retrospective nature, the data being sourced exclusively from one institution, which may limit potential extrapolation to broader populations, and the lack of direct correlation between cytopathology and the definitive gold standard, surgical pathology confirmation.

## Conclusion

In conclusion, our study demonstrates that assessing the AUS rate in the context of PCR, utilizing ThyroSeq v3, serves as a valuable metric for evaluating CP proficiency and overall cytology laboratory performance. Including temporality as a metric can help identify practice trends and provide valuable feedback.

## Data Availability

All data produced in the present work are contained in the manuscript

## Legends

**Supplementary Figure 1.**
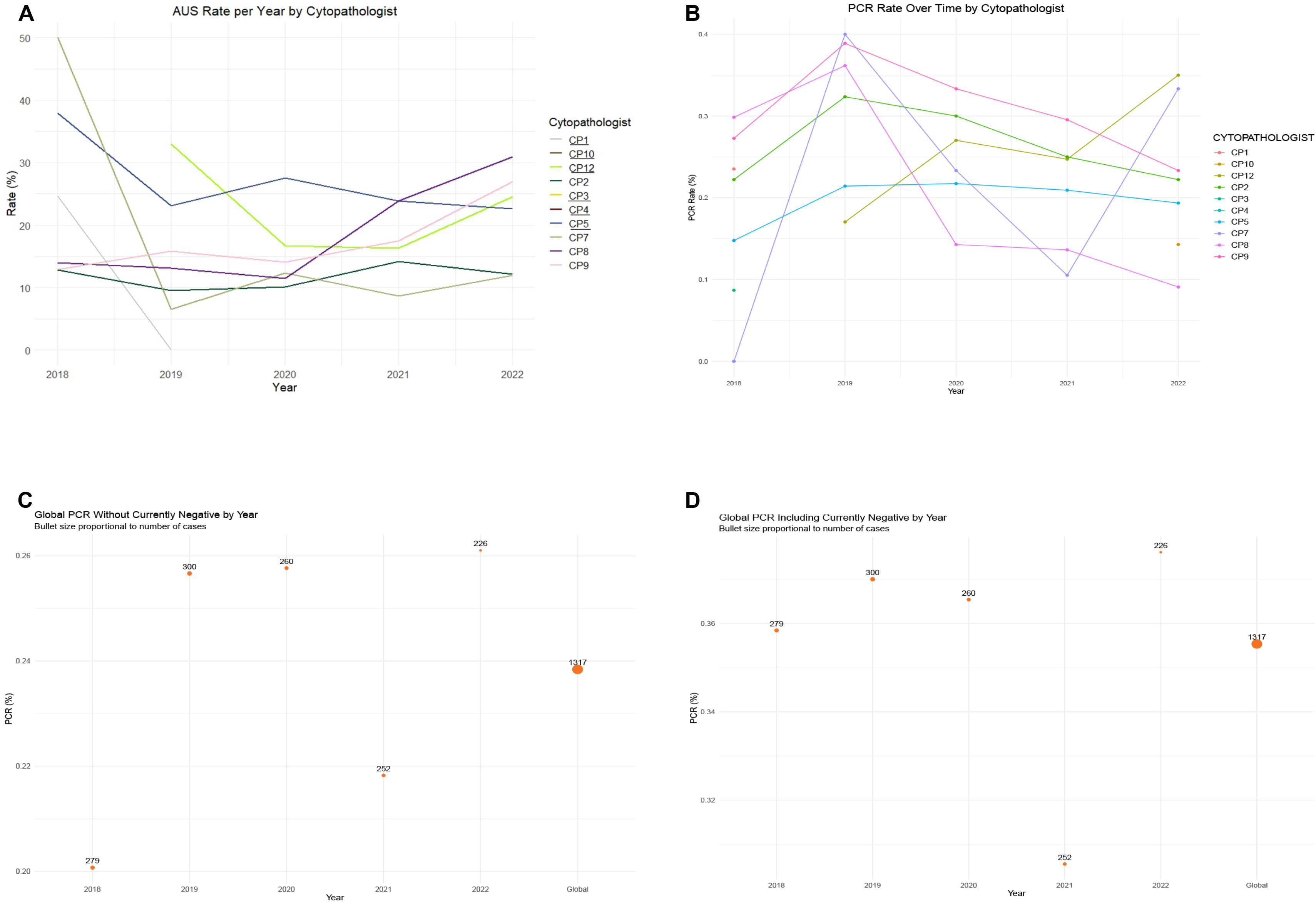
**A)** Line plot showing the AUS rate per cytopathologist per year, similar to figure 4, to compare to the PCR by cytopathologist over time **(B). C)** Scatter plot showing the PCR of the laboratory per year calculated without including *currently negative* cases. **D)** Scatter plot showing the PCR of the laboratory per year calculated including *currently negative* cases. The size of the bullets is proportional to the number of cases, which are depicted atop each bullet.

**Supplementary Figure 2.**
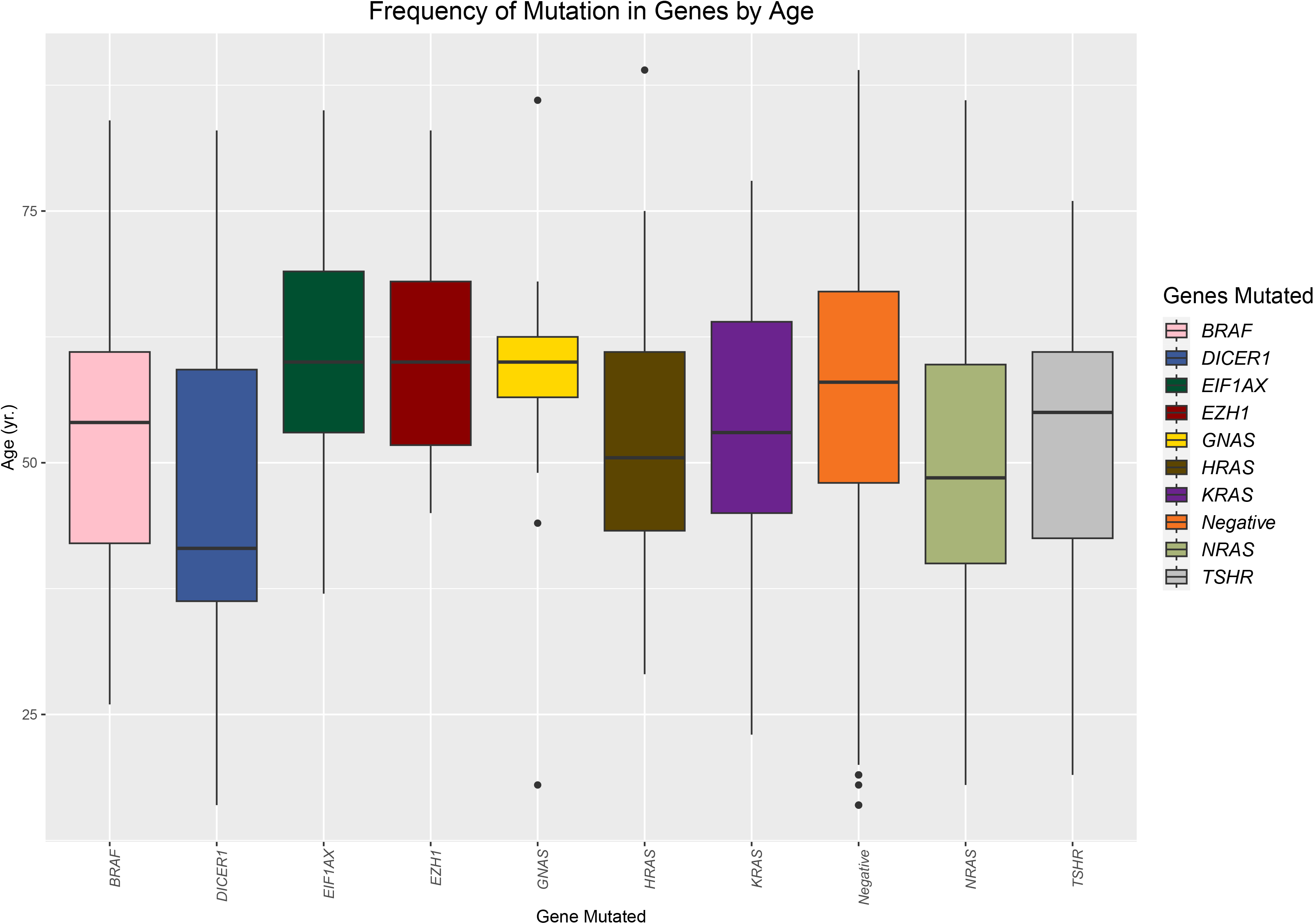
Boxplot showing the frequency of mutations in the most commonly mutated genes based on age. The orange box represents the patients with negative results for comparison.

## References

1. Baloch, Z.W., LiVolsi, V.A., Asa, S.L., Rosai, J., Merino, M.J., Randolph, G., Vielh, P., DeMay, R.M., Sidawy, M.K., and Frable, W.J. (2008). Diagnostic terminology and morphologic criteria for cytologic diagnosis of thyroid lesions: a synopsis of the National Cancer Institute Thyroid Fine-Needle Aspiration State of the Science Conference. Diagn Cytopathol 36, 425–437. 10.1002/dc.20830.

2. Ali, S.Z., Baloch, Z.W., Cochand-Priollet, B., Schmitt, F.C., Vielh, P., and VanderLaan, P.A. (2023). The 2023 Bethesda System for Reporting Thyroid Cytopathology. Thyroid® 33, 1039–1044. 10.1089/thy.2023.0141.

3. Kargi, A.Y., Bustamante, M.P., and Gulec, S. (2017). Genomic Profiling of Thyroid Nodules: Current Role for ThyroSeq Next-Generation Sequencing on Clinical Decision-Making. Mol Imaging Radionucl Ther 26, 24–35. 10.4274/2017.26.suppl.04.

4. VanderLaan, P.A., and Nishino, M. (2022). Molecular testing results as a quality metric for evaluating cytopathologists’ utilization of the atypia of undetermined significance category for thyroid nodule fine-needle aspirations. J Am Soc Cytopathol 11, 67–73. 10.1016/j.jasc.2021.10.001.

5. Cibas, E.S., and Ali, S.Z. (2017). The 2017 Bethesda System for Reporting Thyroid Cytopathology. Thyroid 27, 1341–1346. 10.1089/thy.2017.0500.

6. Ohori, N.P. (2022). A decade into thyroid molecular testing: where do we stand? J Am Soc Cytopathol 11, 59–61. 10.1016/j.jasc.2021.11.002.

7. NCCN Clinical Practice Guideline. Thyroid Carcinoma. Version 4.2023 — August 16, 2023.

8. Martinez Coconubo, D., Levy, J.J., Kerr, D.A., Vaickus, L.J., Vidis, L., Glass, R.E., Gutmann, E.J., Marotti, J.D., and Liu, X. (2023). Use of molecular testing results to analyze the overuse of atypia of undetermined significance in thyroid cytology. Journal of the American Society of Cytopathology. 10.1016/j.jasc.2023.09.002.

9. Ho, A.S., Sarti, E.E., Jain, K.S., Wang, H., Nixon, I.J., Shaha, A.R., Shah, J.P., Kraus, D.H., Ghossein, R., Fish, S.A., et al. (2014). Malignancy Rate in Thyroid Nodules Classified as Bethesda Category III (AUS/FLUS). Thyroid 24, 832–839. 10.1089/thy.2013.0317.

10. Al-Abbadi, M.A., Shareef, S.Q., Yousef, M.M., Almasri, N.M., Mustafa, H.E., Aljawad, H., Ali, J.A., Groves, A., and Alsaihati, Y. (2017). A follow-up study on thyroid aspirates reported as atypia of undetermined significance/follicular lesion of undetermined significance and follicular neoplasm/suspicious for follicular neoplasm: A multicenter study from the Arabian Gulf region. Diagn Cytopathol 45, 983–988. 10.1002/dc.23805.

11. Erivwo, P., and Ghosh, C. (2018). Atypia of Undetermined Significance in Thyroid Fine-Needle Aspirations: Follow-Up and Outcome Experience in Newfoundland, Canada. Acta Cytol 62, 85–92. 10.1159/000486779.

12. Wong, L.Q., LiVolsi, V.A., and Baloch, Z.W. (2014). Diagnosis of atypia/follicular lesion of undetermined significance: An institutional experience. Cytojournal 11, 23. 10.4103/1742-6413.139725.

13. Bernstein, J.M., Shah, M., MacMillan, C., and Freeman, J.L. (2016). Institution-specific risk of papillary thyroid carcinoma in atypia/follicular lesion of undetermined significance. Head Neck 38 Suppl 1, E1210–1215. 10.1002/hed.24193.

14. Hathi, K., Rahmeh, T., Munro, V., Northrup, V., Sherazi, A., and Chin, C.J. (2021). Rate of malignancy for thyroid nodules with AUS/FLUS cytopathology in a tertiary care center - a retrospective cohort study. J Otolaryngol Head Neck Surg 50, 58. 10.1186/s40463-021-00530-0.

15. Wu, H.H.-J., Rose, C., and Elsheikh, T.M. (2012). The Bethesda system for reporting thyroid cytopathology: An experience of 1,382 cases in a community practice setting with the implication for risk of neoplasm and risk of malignancy. Diagn Cytopathol 40, 399–403. 10.1002/dc.21754.

16. Horback, K., Sundling, K.E., Schmidt, R.J., and Cibas, E.S. (2021). Developing dashboards for performance improvement in cytopathology. J Am Soc Cytopathol 10, 535–542. 10.1016/j.jasc.2021.07.001.

17. Silaghi, C.A., Lozovanu, V., Georgescu, C.E., Georgescu, R.D., Susman, S., Năsui, B.A., Dobrean, A., and Silaghi, H. (2021). Thyroseq v3, Afirma GSC, and microRNA Panels Versus Previous Molecular Tests in the Preoperative Diagnosis of Indeterminate Thyroid Nodules: A Systematic Review and Meta-Analysis. Front Endocrinol (Lausanne) 12, 649522. 10.3389/fendo.2021.649522.

18. Steward, D.L., Carty, S.E., Sippel, R.S., Yang, S.P., Sosa, J.A., Sipos, J.A., Figge, J.J., Mandel, S., Haugen, B.R., Burman, K.D., et al. (2019). Performance of a Multigene Genomic Classifier in Thyroid Nodules With Indeterminate Cytology: A Prospective Blinded Multicenter Study. JAMA Oncology 5, 204–212. 10.1001/jamaoncol.2018.4616.

19. Desai, D., Lepe, M., Baloch, Z.W., and Mandel, S.J. (2021). ThyroSeq v3 for Bethesda III and IV: An institutional experience. Cancer Cytopathology 129, 164–170. 10.1002/cncy.22362.

20. Gajzer, D.C., Tjendra, Y., Kerr, D.A., Algashaamy, K., Zuo, Y., Menendez, S.G., Jorda, M., Garcia-Buitrago, M., Gomez-Fernandez, C., and Velez Torres, J.M. (2022). Probability of malignancy as determined by ThyroSeq v3 genomic classifier varies according to the subtype of atypia. Cancer Cytopathology 130, 881–890. 10.1002/cncy.22617.

21. Hong, S., Lee, H., Cho, M.-S., Lee, J.E., Sung, Y.-A., and Hong, Y.S. (2018). Malignancy Risk and Related Factors of Atypia of Undetermined Significance/Follicular Lesion of Undetermined Significance in Thyroid Fine Needle Aspiration. Int J Endocrinol 2018, 4521984. 10.1155/2018/4521984.

22. Ohori, N.P., Landau, M.S., Manroa, P., Schoedel, K.E., and Seethala, R.R. (2020). Molecular-derived estimation of risk of malignancy for indeterminate thyroid cytology diagnoses. J Am Soc Cytopathol 9, 213–220. 10.1016/j.jasc.2020.03.004.

23. Chiosea, S., Hodak, S.P., Yip, L., Abraham, D., Baldwin, C., Baloch, Z., Gulec, S.A., Hannoush, Z.C., Haugen, B.R., Joseph, L., et al. (2023). Molecular Profiling of 50 734 Bethesda III-VI Thyroid Nodules by ThyroSeq v3: Implications for Personalized Management. The Journal of Clinical Endocrinology & Metabolism, dgad220. 10.1210/clinem/dgad220.

24. Mon, S.Y., Riedlinger, G., Abbott, C.E., Seethala, R., Ohori, N.P., Nikiforova, M.N., Nikiforov, Y.E., and Hodak, S.P. (2018). Cancer risk and clinicopathological characteristics of thyroid nodules harboring thyroid-stimulating hormone receptor gene mutations. Diagn Cytopathol 46, 369–377. 10.1002/dc.23915.

